# Height-age as an alternative to height-for-age z-scores to assess the effect of interventions on child linear growth in low- and middle-income countries

**DOI:** 10.1101/2024.07.15.24310427

**Authors:** Kelly M Watson, Alison SB Dasiewicz, Diego G Bassani, Chun-Yuan Chen, Huma Qamar, Karen M O’Callaghan, Daniel E Roth

## Abstract

**Background:** Assessments of the efficacy of interventions to improve child growth are often based on differences in mean height-for-age z-scores (HAZ) and stunting (HAZ<-2) in randomized controlled trials (RCTs). However, this approach does not account for children’s starting skeletal age and does not enable assessment of the extent to which interventions optimized linear growth.

**Objective:** To develop and apply a new method using height-age to express linear growth effects in RCTs.

**Methods:** Longitudinal individual participant data (IPD) from a Bangladeshi trial cohort were used to compare height-age estimates derived from individual-level heights, mean raw height, or mean HAZ. Then, using average height-age as a proxy for skeletal age, we developed the ‘proportion of maximal benefit’ (PMB) metric to quantify intervention effects relative to optimal growth for children’s starting skeletal age. Optimal growth occurs when height-age increases in parallel with chronological age (i.e., PMB=100%) whereas no effect (versus control) corresponds to a PMB of 0%. Linear growth outcomes in 4 published RCTs of nutrition-specific interventions were re-expressed as mean height-age and PMB, and compared to effects conventionally expressed as intervention-versus-control mean differences (MD) in HAZ.

**Results:** Mean height-age could be derived from any published estimate of mean raw height or mean HAZ; however, to calculate the PMB, height or HAZ data were required at both the beginning and end of the observation period. Interpretations of intervention effects were consistent when expressed as either the height-age MD or HAZ MD. In contrast, the PMB does not have a corresponding metric on the HAZ scale, and therefore provided a new way to quantify intervention efficacy.

**Conclusion:** Height-age can be used as an alternative to HAZ to express intervention effects. The PMB has the advantage of conveying the extent to which an intervention improved average linear growth in relation to a biologically-defined benchmark.

## INTRODUCTION

Child height is a general marker of health and nutritional status and is commonly used as an outcome in interventional trials and evaluations of programs aimed at reducing undernutrition in low– and middle-income countries (LMICs) (1, 2). Sex– and age-standardized height-for-age z-scores (HAZ) and stunting (HAZ<-2) relative to the WHO Growth Standards (WHO-GS) are conventionally used to quantify growth deficits in individuals and populations (3). Candidate interventions to improve child health and nutrition are often considered for adoption as public health programs in LMICs based on the extent to which they increase mean HAZ and reduce stunting prevalence in randomized controlled trials (RCTs). However, the conventional reliance on HAZ and stunting to assess changes in child health and nutritional status have been questioned (4–8). One limitation is that the use of stunting may lead to a disproportionate focus on the sub-group of children classified as stunted, even though linear growth faltering affects the entire height-for-age distribution in LMICs (5, 6). Another concern is that it is unrealistic to expect a large change in HAZ or stunting in response to a single, short-term intervention given that the cause of linear growth faltering is multi-factorial, leading to interventions being perceived as failures even if they are shown to have other health or social benefits (4, 8). Additionaly, the use of HAZ (or raw height) in linear growth outcome analyses does not enable definition of a population’s maximum biologically plausible growth that could occur over an observation period, so there is no objective benchmark to evaluate the adequacy of a response to an intervention using the HAZ or raw height scale.

Height-age, defined as the age at which the observed height of children would be considered normal, is an alternative way of expressing linear growth that conveys information about the stage of maturation of long bones (‘skeletal age’). Skeletal maturation is driven by mechanisms intrinsic to the growth plate, such that skeletal age and chronological age become uncoupled (skeletal age < chronological age) under growth-inhibiting conditions (e.g., nutritional deficiencies) (9–11). Following remediation of the causes of growth impairments, linear growth has been observed to proceed at a rate expected for the younger skeletal age (or height-age), rather than chronological age (9, 12–15).

Setting expectations for successful interventions based on HAZ and stunting, which are normalized relative to expected growth for children’s chronological age, may contribute to the perception that candidate interventions often fail to improve linear growth (4, 8). The aim of the present methodological study was to develop metrics to use height-age as an alternative outcome to stunting and HAZ in RCT analysis and to demonstrate these metrics in a pilot study to enable their wider application and further validation in future research. As this was a novel application of height-age, we first compared different ways of deriving height-age estimates using RCT datasets, which enabled us to propose a preferred method for future uses. We also developed a new metric based on height-age that quantifies the extent of linear growth improvement achieved by interventions on the height-age scale.

## METHODS

### Data sources

Individual participant data (IPD) from the Maternal Vitamin D for Infant Growth (MDIG) trial (16) and follow-up BONe and mUScle health in Kids (BONUSKids) study in Dhaka, Bangladesh (17) were used to compare height-age estimates generated from different expressions of height data. Briefly, the MDIG trial was a randomized, placebo-controlled, dose-ranging trial of maternal vitamin D supplementation during pregnancy and lactation. One group received neither prenatal or postpartum vitamin D (placebo group); three groups received prenatal supplementation only (4200 IU, 16,800 IU, or 28,000 IU per week); one group received prenatal and postpartum supplementation up to 26-weeks (28,000 IU per week; denoted as ‘28000:28000 IU’ which corresponds to ‘prenatal:postpartum’). The MDIG trial assessed linear growth in children from birth to 24 months of age, and the BONUSKids follow-up study assessed the MDIG trial cohort at 4 years of age. Ethical approval was obtained from The Hospital for Sick Children’s (SickKids) research ethics board (REB) for use of MDIG and BONUSKids IPD in this study (SickKids REB #1000079659). Published estimates from selected intervention trials (18–22) based on inclusion criteria (outlined below) were used in a pilot application of novel height-age-based methods.

### Existing height-age derivation methods

Height-age was defined as the age at which the WHO-GS median length or height equaled children’s observed mean length or height (hereafter, “height” will refer to measures of length or height). The WHO-GS height-for-age curves, which include the median growth trajectory, are generated by the lamda-mu-sigma (LMS) method (23), using the publicly available WHO-LMS table containing the data underlying the WHO-GS curves (‘lenanthro.dta’ file) (24). The relevant parameters from the WHO-GS LMS table for calculating height-age were the median height (cm), and the corresponding age (in days) and sex (male/female). Height-age is the age corresponding to the median height in the WHO-GS LMS table closest to the observed mean height. If group-level mean HAZ was the only estimate available in the published literature (e.g., only mean HAZ, not mean height, was reported), the observed mean height was back-calculated from the reported mean HAZ. Equation 1 was rearranged to solve for ‘y’ (the observed height) (3). The median height (M) and coefficient of variation (S), corresponding to the average age of children at the time (t) mean HAZ was measured, were selected from the WHO-LMS table to facilitate this calculation. The power (L) in the WHO-LMS table is always equal to 1 (24).

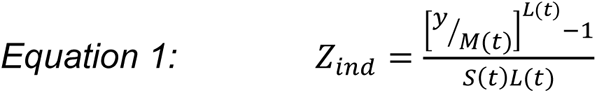

The Supplementary Data file from Mansukoski *et al*. includes publicly available code to derive height-age from mean HAZ (25).

### Extension of height-age derivation methods

We considered mean height (rather than individual-level height or mean HAZ) to be the preferred expression of height data for deriving height-age, since the expectation that growth tracks along the median trajectory is applicable at the population level but not for individual children, and mean height can be directly compared to the WHO-GS median (avoiding the back-calculation required for HAZ). Building on prior work, we documented a method to derive height-age from mean height when data is disaggregated by sex, or combining sexes using a sex-weighted or simple average of the WHO-GS LMS table parameters (26).

We developed a new workflow for calculating height-age when children were measured at or near two years of age (**Figure 1**). At this age, there is typically a switch from measuring supine (recumbent) length to standing height which leads to a slight reduction in measured stature (∼0.7cm) (27). The WHO-LMS table presents the median as recumbent length when age is <731 days (2 years) and standing height when age is >=731 days (24). The WHO macro for calculating z-scores converts height measurements to length for children under 2 years of age (by adding 0.7cm), or length to height for children >=2 years (by subtracting 0.7cm) (28). Figure 1 shows how we adjusted the WHO median values presented in the LMS table to ensure fair comparison with observed measurements when determining height-age. For example, if using an observed mean height measurement of 2-year-old children who have experienced prior linear growth faltering to calculate height-age, it can be assumed that their calculated average height-age will be *less* than 2 years. It is then necessary to convert WHO-GS median values for children <2 years of age (which typically represent supine length measurements) to standing height measurements, by subtracting 0.7cm from all medians representing supine length measurements. This ensures the observed height is compared with a median value reflecting a standing height measurement.

**FIGURE 1.**
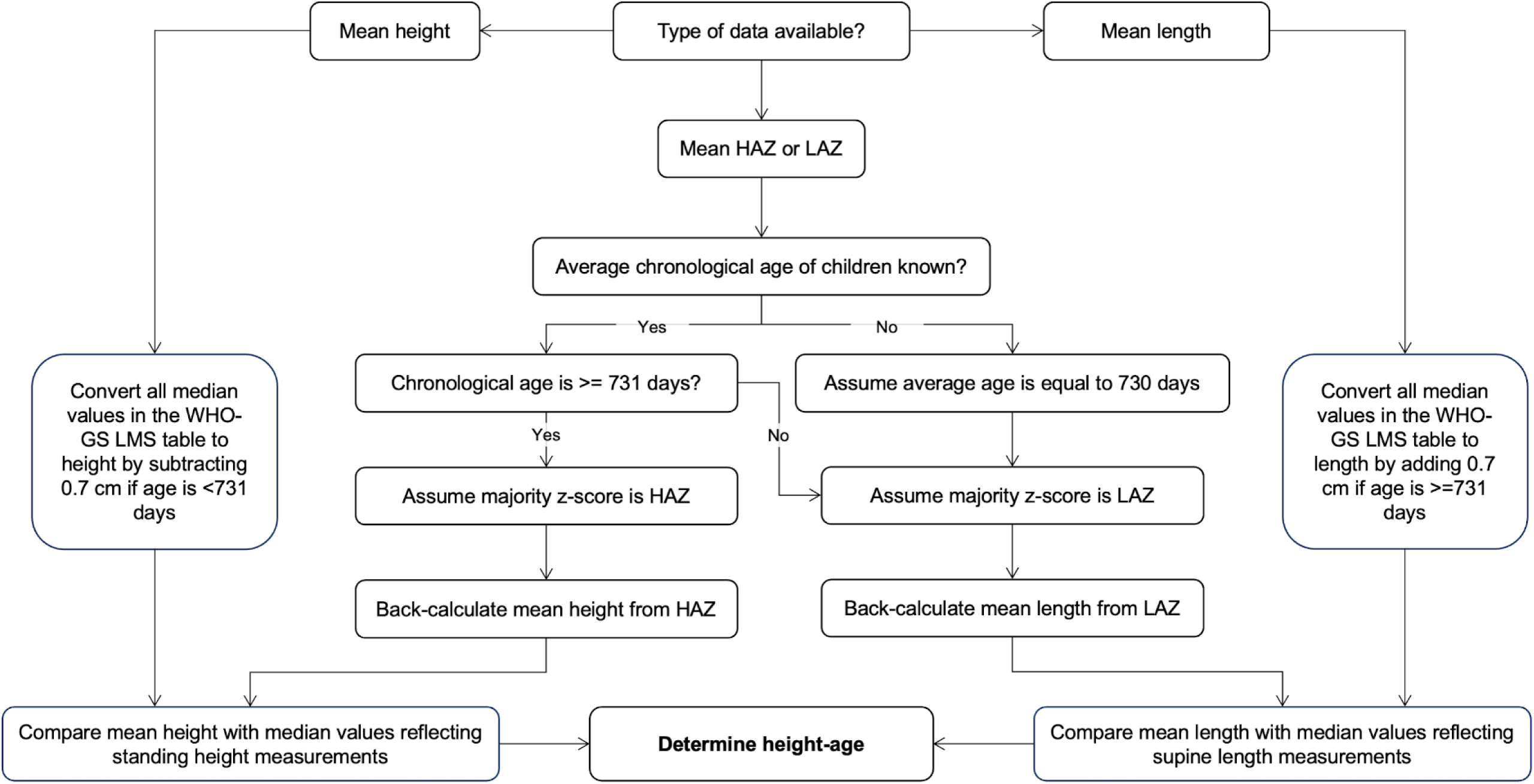
Workflow to determine height-age of children measured at or near two years of age. This workflow is intended to be used for studies in which a scheduled follow-up was at, or near, 2 years of age, as it addresses the 0.7 cm discontinuity in the WHO-GS median length/height values before and after 2 years (731 days) of age (due to the transition from supine length to standing height measurement). “Assume majority z-score is HAZ” and “Assume majority z-score is LAZ” indicates the need to assume that this was the type of z-score that was calculated for a majority of children by the WHO macro for calculating z-scores. It is then assumed that either a standing height (for majority HAZ) or recumbent length (for majority LAZ) measurement is back-calculated from the z-score. This affects the subsequent adjustment made to the WHO-GS median values to ensure fair comparison between the observed height or length and reference values (WHO-GS median) when estimating height-age. HAZ, height-for-age Z-score; LAZ, length-for-age Z-score; LMS, lambda-mu-sigma; WHO-GS, World Health Organization Growth Standard.

### Development of the proportion of maximal benefit (the ‘PMB’) metric

The ‘proportion of maximal benefit’ (PMB) was developed to quantify the extent to which children in an intervention group reached a theoretical benchmark of optimal growth, defined as the normal growth rate for their baseline height-age. The normal (‘optimal’) growth rate for children’s average starting height-age was defined by the median trajectory of the WHO-GS height-for-age chart; however, as the expected growth rate constantly changes on the height-for-age scale (**Figure 2 Panel A**), the median trajectory was converted to a plot of *height-age*-for-age, by changing the y-axis from height (cm) to height-age (days) (**Figure 2 Panel B**). On this new scale, the normal growth rate is achieved when height-age increases the same amount as chronological age (slope = 1), and in contrast to the height-for-age curve, this relationship is constant across age (straight-line function).

**FIGURE 2.**
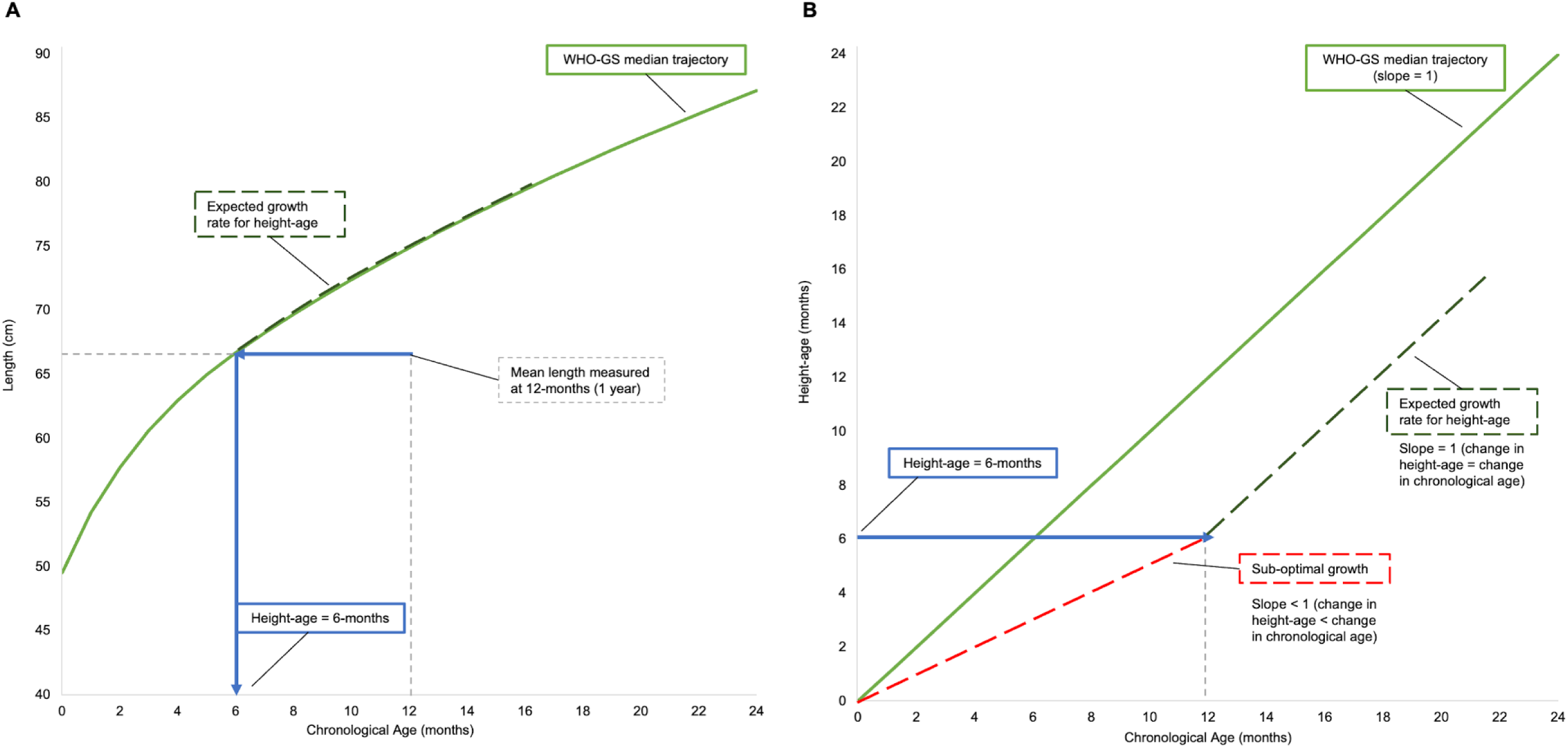
Schematic representation of the correspondence between the expected growth rate for baseline height-age given A) a conventional length-for-age curve and B) a plot of height-age as a function of chronological age. Assuming a hypothetical group of 1-year-old children who have experienced linear growth faltering, such that the average height-age is <1 year, were measured at the start of an intervention, panel A) shows that the expected growth rate for children’s baseline height-age changes at any given age on the length-for-age scale, whereas panel B) shows that the expected growth rate on the height-age-for-age scale is a constant relationship across ages (slope = 1). WHO-GS, World Health Organization Growth Standard. Data source: WHO. WHO Child Growth Standards STATA igrowup package [Internet]. 2022 [cited 2023 Jul 19]. Available from: https://github.com/unicef-drp/igrowup_update

Using this simple 1-to-1 expected relationship between height-age and chronological age, the theoretical maximum growth potential of children in the intervention group can be defined by the group’s change in chronological age during the observation period (ΔCA_I_). The observed effect of the intervention, expressed as the intervention group’s change in height-age during the observation period (ΔHA_I_), can be compared to this benchmark. The control group’s change in height-age during the observation period (ΔHA_C_) was used as the lower bound, as it was assumed that if the intervention provided no benefit at all, growth in the intervention group(s) would proceed at the same rate as in the control group. All changes (Δs) were measured over the same observation period, expressed in the same units of time (days, months, etc.), and input into *Equation 2* to calculate the PMB.

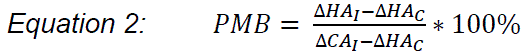

A PMB of 0% means that the change in height-age was equal in the intervention and control groups during the intervention period, implying that the intervention had no effect. A PMB of 100% means that height-age in the intervention group increased at the same rate as chronological age, which implies achievement of the optimal growth rate (Figure 2). While achieving 100% is mathematically possible, it is expected that the PMB resulting from a single, short-term intervention that addresses only a fraction of the factors inhibiting linear growth will be much lower than 100%. Therefore, achieving a PMB of 100% should not be considered a realistic target for successful interventions in such settings; however, further use of the PMB for a variety of interventions will be required to establish specific PMB thresholds for policy-relevant applications.

### Application of height-age and the PMB in a pilot study

#### i) Inclusion criteria

Inclusion criteria for intervention trials included in this pilot study were: 1) RCT or a follow-up report of a previously conducted RCT; 2) conducted in a low, lower-middle, or upper-middle income country as defined by the World Bank classifications (29); 3) study population was infants or young children (between 0 to 5 years of age); 4) intervention was nutrition-specific (e.g., vitamin D supplementation); 5) study or group-level HAZ at baseline and end-line of the intervention period were available.

The MDIG trial and BONUSKids study were also included and summary estimates were generated to emulate having access to study or group-level data only, as was the case with the other studies included. As well, for comparability with another vitamin D supplementation trial included, only the 28000:28000 IU/week maternal vitamin D supplementation group was included (the only group with postpartum supplementation), the intervention period was defined as 0 to 6 months of age (the time post-delivery in which the maternal intervention was ongoing), and the post-intervention period as 6 to 48 months.

#### ii) Preliminary steps

To re-express trial outcomes, mean HAZ was used to derive mean height-age as it was more commonly reported than mean height and thereby enabled a more consistent approach across included trials. As mean height was *a priori* defined as the preferred method for deriving mean height-age, we compared height-age estimates (means and 95% CIs) derived from mean HAZ to corresponding estimates derived from mean height or individual-level heights. The purpose of this preliminary step was to ensure mean HAZ-derived height-age estimates in the pilot study were similar to estimates derived from preferred methods. Individual-level child data (age, sex, length or height) from the MDIG trial and BONUSKids study at the 3, 6, 9, 12, 24, and 48 months encounters were used to generate these height-age estimates from the different expressions of height data (**Figure 3**). Differences between mean height-age estimates derived from mean HAZ versus mean height, and height-age standard errors (SEs) derived from mean HAZ versus individual-level heights were calculated. The mean, median, standard deviation (SD), kurtosis, and skewness generated from kernel density plots of individual-level height, HAZ, and height-age were also compared at the selected timepoints using MDIG and BONUSKids data to identify any statistical differences of the height-age distribution that may influence interpretation.

**FIGURE 3.**
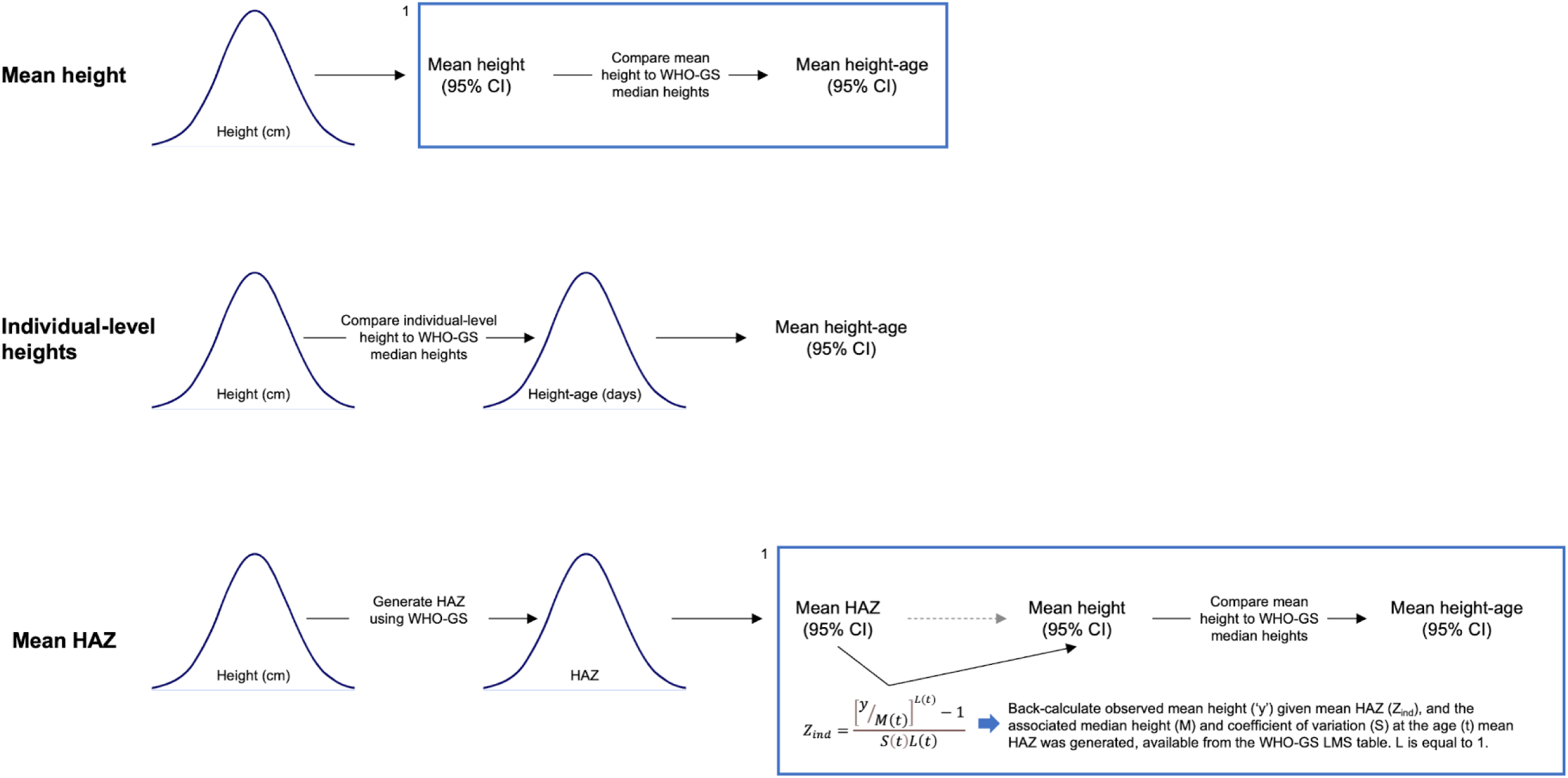
Approaches to derive height-age estimates based on available data. ‘Mean height’ represents using mean height or length to derive height-age; ‘Individual-level heights’ represents using individual-level heights or lengths to derive individual-level height-ages; ‘Mean HAZ’ represents using mean HAZ or LAZ to derive height-age. The normal distributions are used to depict individual-level child data from the MDIG trial and BONUSKids study. ^1^The steps within the box represent the assumption that only mean height or HAZ are available for deriving height-age; the preceding steps are included to show how mean height and HAZ were generated from individual-level data. CI, confidence interval; HAZ, height-for-age z-score; WHO-GS, World Health Organization Growth Standard.

#### iii) Data extraction from included trials in the pilot study

Mean HAZ reported at baseline and end-line were extracted from published reports of included trials. The average age of children at the time of measurement was extracted; if the precise average age at baseline and/or end-line was not reported, the average age was assumed to equal the timing of measurement (e.g., if scheduled end-line follow-up was at 6 months, the average age was assumed to equal baseline age plus 6 months). Analyses were conducted combining sexes as it was assumed this was the manner in which data would be most often reported. If the ratio of females and males was reported, this was extracted to generate a sex-weighted WHO-GS LMS table; otherwise, an equal ratio was assumed.

#### iv) Statistical analyses – application of the PMB and comparator outcomes

Extracted data were used to re-express mean HAZ and the associated 95% CI as mean height-age with 95% CIs. The height-age mean difference (MD) (intervention minus control) at the study end-line was determined with 95% CIs (30). This was compared to how results were reported in the published paper (HAZ MD); to ensure direct comparability, we recalculated the HAZ MD using the same method as used for the height-age MD (30). The MD between groups’ changes in height-age during the observation period (end-line height-age minus baseline height-age) was also determined with 95% CIs using methods outlined in **Supplemental Methods** and **Supplemental Table 1**, as an intermediary step to facilitate calculation of the PMB. To calculate 95% CIs for the PMB, the 95% confidence bounds for the MD between groups’ changes in height-age were substituted in the PMB numerator in *Equation 2*, as the numerator in *Equation 2* is the MD between the intervention and control group’s change in height-age over the observation period (i.e., to calculate the upper 95% bound for the PMB, the upper bound of the 95% CI for the MD between groups’ changes in height-age was substituted into the numerator of the PMB metric; the same was done to calculate the lower bound). As a sensitivity analysis, we repeated this exercise using mean height if it was also reported at baseline and/or end-line to compare height-age and PMB estimates generated from mean height versus mean HAZ. All analyses were performed in STATA software version 17 (STATA Corp LLC).

## RESULTS

Across six age timepoints in the MDIG/BONUSKids cohort, the maximum absolute difference between mean height-age estimates generated from mean HAZ versus mean height was 1 day (Range: 0 to 1). The maximum difference between SEs generated from mean HAZ versus from a distribution of height-ages (generated from individual-level heights), was 0.3 day (Range: 0 to 0.3) (**Table 1**). These results were similar when the analyses was disaggregated by sex. The distribution of individual height-age estimates consistently showed slightly greater skewing (Mean skewness: 0.4 for height-age; 0.05 for height; 0.01 for HAZ) and excess kurtosis (Mean kurtosis: 3.5 for height-age; 3.2 for height; 3.1 for HAZ) compared to the raw height and HAZ distributions (**Table 2**).

**TABLE 1.** Height-age generated from mean length or height (’Mean Height’), mean LAZ or HAZ (‘Mean HAZ’), or individual-level lengths or heights (‘Individual-level Heights’), using individual participant data from the Maternal vitamin D for Infant Growth (MDIG) trial and the BONe and mUScle health in Kids (BONUSKids) follow-up the of the MDIG cohort.

| Encounter<sup>1</sup><br>(months) | *n*<br>participants | Mean age, d<br>(Min, Max) | Height-age, d |  |  |  |  |  |
| --- | --- | --- | --- | --- | --- | --- | --- | --- |
|  |  |  | Mean Height<sup>2</sup> |  | Individual-level Heights<sup>2</sup> |  | Mean HAZ<sup>3</sup> |  |
|  |  |  | Mean (95% CI) | SE | Mean (95% CI) | SE | Mean (95% CI) | SE |
| 3 | 1028 | 94 (91, 162) | 73 (72, 74) | 0.5 | 74 (73, 76) | 0.8 | 73 (72, 75) | 0.8 |
| 6 | 1037 | 185 (182, 262) | 151 (149, 154) | 1.3 | 154 (152, 157) | 1.3 | 152 (149, 154) | 1.3 |
| 9 | 1128 | 277 (273, 348) | 233 (230, 236) | 1.5 | 235 (232, 239) | 1.6 | 233 (230, 236) | 1.5 |
| 12 | 1165 | 368 (364, 427) | 307 (304, 311) | 1.8 | 310 (306, 313) | 1.9 | 307 (304, 311) | 1.8 |
| 24 | 1100 | 733 (666, 807) | 592 (586, 599) | 3.3 | 597 (591, 603) | 3.2 | 592 (586, 599) | 3.3 |
| 48 | 640 | 1510 (1431, 1553) | 1242 (1226, 1259) | 8.4 | 1251 (1235, 1268) | 8.5 | 1242 (1226, 1258) | 8.2 |

<sup>1</sup>3-, 6-, 9-, 12-, and 24-month encounters measured during the MDIG trial; 48-month encounter measured during the BONUSKids study. <sup>2</sup>Length (mean or individual-level) used at the 3-, 6-, 9-, 12-, and 24-month encounters; height (mean or individual-level) used at the 48-month encounter.
<sup>3</sup>Mean LAZ used at the 3-, 6-, 9-, 12-month encounters; mean HAZ used at the 24- and 48-month encounters.
CI, confidence interval; d, days; HAZ, height-for-age z-score; IPD, individual participant data; LAZ, length-for-age z-score; SE, standard error.

**TABLE 2.** Statistical properties of the height, HAZ, and height-age distributions, generated using individual participant data from the Maternal vitamin D for Infant Growth (MDIG) trial and the BONe and mUScle health in Kids (BONUSKids) follow-up the of the MDIG trial cohort.

| Encounter<sup>1</sup><br>(months) | Length or height<sup>2</sup> (cm) |  |  |  |  | LAZ or HAZ<sup>2</sup> |  |  |  |  | Height-age<sup>3</sup> (months) |  |  |  |  |
| --- | --- | --- | --- | --- | --- | --- | --- | --- | --- | --- | --- | --- | --- | --- | --- |
|  | Mean | Med. | SD | Kurt. | Skew. | Mean | Med. | SD | Kurt. | Skew. | Mean | Med. | SD | Kurt. | Skew. |
| 3 | 59.0 | 59.0 | 2.0 | 3.6 | 0.1 | -0.9 | -0.9 | 0.9 | 3.1 | -0.2 | 2.5 | 2.4 | 0.7 | 5.4 | 0.7 |
| 6 | 64.9 | 65.0 | 2.2 | 3.0 | 0.0 | -0.9 | -0.8 | 1.0 | 3.0 | -0.1 | 5.1 | 5.0 | 1.2 | 3.1 | 0.4 |
| 9 | 69.2 | 69.2 | 2.4 | 3.1 | -0.1 | -0.9 | -0.9 | 1.0 | 3.1 | -0.1 | 7.7 | 7.7 | 1.7 | 3.0 | 0.2 |
| 12 | 72.5 | 72.6 | 2.6 | 3.0 | -0.1 | -1.0 | -1.0 | 1.0 | 3.1 | -0.1 | 10.2 | 10.1 | 2.0 | 3.0 | 0.2 |
| 24 | 82.9 | 82.8 | 3.4 | 3.2 | 0.1 | -1.4 | -1.4 | 1.1 | 3.2 | 0.1 | 19.7 | 19.3 | 3.5 | 3.3 | 0.4 |
| 48 | 98.7 | 98.5 | 4.4 | 3.2 | 0.3 | -1.2 | -1.3 | 1.0 | 3.2 | 0.3 | 40.8 | 40.2 | 7.1 | 3.2 | 0.5 |

<sup>1</sup>3-, 6-, 9-, 12-, and 24-month encounters measured during the MDIG trial; 48-month encounter measured during the BONUSKids study. <sup>2</sup>Length and LAZ at the 3-, 6-, 9-, 12-, and 24-month encounters; height and HAZ at the 48-month encounter. <sup>3</sup>Height-age generated at the individual-level, from length (3-, 6-, 9-, 12-, and 24-month encounters) or height (48-month encounter).
HAZ, height-for-age z-score; Kurt., kurtosis; LAZ, length-for-age z-score; Med., median; SD, standard deviation; Skew., skewness.

Including MDIG and BONUSKids, 7 reports of 4 trials of 2 distinct nutrition-specific interventions implemented in LMICs were included in the pilot study (**Table 3**). The interventions were the provision of 1 egg/day from 6 to 12 months of age (the Lulun and Mazira trials) and maternal or infant vitamin D supplementation (the Delhi Infant Vitamin D Supplementation (DIVIDS) and MDIG trials). Group/study-level mean HAZ was available at baseline, end-line, and post-intervention follow-up (if applicable) for all included studies. Group-level baseline LAZ for the DIVIDS trial was obtained from another report (31).

**TABLE 3.** Characteristics of trials and available follow-up studies included in the pilot study.

| Country, year(s) of study, study name, *n*, authors | Intervention | Effect on linear growth | Authors' conclusions |
| --- | --- | --- | --- |
| Ecuador, 2015, Lulun, *n*=163, Iannotti et al. (19) | Infants provided 1 egg/day from age 6-12 months | LAZ MD: 0.63 (95% CI: 0.38, 0.88) | Egg intervention significantly increases linear growth |
| Ecuador, 2017, Lulun follow-up, *n*=135, Iannotti et al. (18) | Follow-up of Lulun trial cohort after 2 years | HAZ MD: 0.082 (95% CI: -0.15, 0.31) | Results from Lulun trial attenuated |
| Malawi, 2018-2019, Mazira Project, *n*=660, Stewart et al. (20) | Infants provided 1 egg/day from age 6-12 months | LAZ MD: 0.07 (95% CI: -0.01, 0.15)<sup>1</sup> | Egg intervention did not benefit linear growth |
| India, 2007-2010, DIVIDS, *n*=2079, Kumar et al. (21) | Weekly infant vitamin D supplementation (1400 IU/week) from birth to 6 months | LAZ MD: 0.12 (95% CI: 0.02, 0.21)<sup>2</sup> | Vitamin D supplementation significantly increased linear growth |
| India, 2012-2014, DIVIDS follow-up, *n*=912, Kumar et al. (22) | Follow-up of DIVIDS trial cohort after 3-6 years | HAZ MD: 0.06 (95% CI: -0.07, 0.19)<sup>3</sup> | Results from DIVIDS attenuated |
| Bangladesh, 2014-2018, MDIG, *n*=1300, Roth et al. (16) | Varying doses<sup>4</sup> of maternal prenatal and postpartum weekly vitamin D supplementation | No significant difference between groups LAZ at 1 year (*p*=0.25)<sup>5</sup> | Maternal vitamin D supplementation does not improve infant's linear growth |
| Bangladesh, 2018-2020,<br>BONUSKids, $n=642$,<br>O'Callaghan et al. (17) | Follow-up of MDIG trial cohort<br>at 4 years | No significant difference<br>between groups HAZ at 4<br>years ($p=0.81$)<sup>5</sup> | No comment on children's<br>linear growth |

<sup>1</sup>Minimally adjusted comparison reported in Stewart et al. shown here; interpretation of effect unchanged when fully adjusted comparison considered. <sup>2</sup>Adjusted comparison reported in Kumar et al. (DIVIDS) shown here; effect is insignificant when unadjusted comparison considered. <sup>3</sup>Unadjusted comparison reported in Kumar et al. (DIVIDS follow-up) shown here; interpretation of effect unchanged when fully adjusted comparison considered. <sup>4</sup>One group received neither prenatal or postpartum vitamin D (placebo group); three groups received prenatal supplementation only (4200 IU, 16,800 IU, or 28,000 IU per week); one group received prenatal and postpartum supplementation up to 26-weeks (28,000 IU/week). <sup>5</sup>Analysis of variance (ANOVA) for multiple group comparisons used to measure effect on linear growth in Roth et al. (MDIG) and O'Callaghan et al. (BONUSKids).
BONUSKids, BONE and mUScle health in Kids; CI, confidence interval; DIVIDS, Delhi Infant Vitamin D Supplementation; HAZ, height-for-age z-score; LAZ, length-for-age z-score; MD, mean difference; MDIG, Maternal Vitamin D for Infant Growth.

The egg intervention introduced in the Lulun trial led to a significant increase in mean LAZ at 12 months (19), while the same intervention in the Mazira trial did not (20). Infant vitamin D supplementation in the DIVIDS trial led to a significant increase in LAZ at 6 months (21), while maternal vitamin D supplementation in the MDIG trial did not improve children’s linear growth at 1 year (16). Three of the four trials had post-intervention follow-ups (Lulun, DIVIDS, and MDIG), and no sustained or latent improvements in linear growth were observed (17, 18, 22).

Inferences and directions of the intervention effects at end-line were consistent when expressed on the HAZ and height-age scales (**Table 4**). For example, the Mazira trial results expressed as the LAZ or height-age MD both indicated that the intervention did not significantly improve linear growth. However, neither of these effect estimates conveyed the extent of linear growth improvement achieved by this intervention relative to a benchmark of children’s optimal growth potential. The PMB revealed that the provision of 1 egg/day for 6 months in this population achieved 9.4% of the intervention group’s growth potential beyond growth in the control group. For the Lulun trial, the LAZ and height-age MD both showed that this intervention improved growth; the PMB suggested that this intervention achieved 94% of the intervention group’s optimal growth potential. For the DIVIDS and MDIG trials, height-age was not derived at baseline as the intervention began at birth, so the PMB could not be calculated (an intermediate step to enable calculation of the PMB is determining the change in height-age from baseline to end-line). For the trials with post-intervention follow-ups, the PMBs ranged from –0.28% to 5.2% (Table 4).

**TABLE 4.** Effect of interventions quantified using conventional and height-age-based methods.

| Trial (intervention duration) | Time of assessment<sup>2</sup> | Effect estimate (95% CI) |  |  |
| --- | --- | --- | --- | --- |
|  |  | Mean difference (intervention - control) |  | PMB, % |
|  |  | LAZ or HAZ<sup>3</sup> | Height-age, d |  |
| MDIG<sup>1</sup> (0-6 months) | End of trial | 0.11 (-0.077, 0.30) | 4 (-3, 11) | NA<sup>4</sup> |
|  | Follow-up | 0.06 (-0.20, 0.32) | 16 (-38, 70) | 5.2 (-15, 25) |
| DIVIDS (0-6 months) | End of trial | 0.11 (0.001, 0.22) | 4 (0, 8) | NA<sup>4</sup> |
|  | Follow-up | 0.06 (-0.07, 0.19) | 14 (-16, 44) | 2.5 (-4.3, 9.3) |
| Lulun (6-12 months) | End of trial | 0.32 (-0.067, 0.71) | 12 (-11, 35) | 94 (50, 138) |
|  | Follow-up | -0.09 (-0.40, 0.22) | -40 (-96, 16) | -0.28 (-0.51, -0.06) |
| Mazira (6-12 months) | End of trial | 0.02 (-0.15, 0.19) | 4 (-7, 15) | 9.4 (-9.4, 28) |

<sup>1</sup>Different primary end-line selected for this study (6 months) than the primary end-line reported in the MDIG trial (1 year). <sup>2</sup>'Follow-up' represents measurement of children after an elapsed period of time following cessation of the trial. <sup>3</sup>LAZ mean difference (MD) for all trials at end of trial; HAZ MD for all trials with follow-up. <sup>4</sup>Height-age could not be calculated at baseline because methods were not developed in this work to address cases when the age of children approximates 0 days.
CI, confidence interval; d, days; DIVIDS, Delhi Infant Vitamin D Supplementation; HAZ, height-for-age z-score; LAZ, length-for-age z-score; MDIG, Maternal Vitamin D for Infant Growth; PMB, proportion of maximal benefit.

The Lulun and Mazira trial results were plotted as LAZ-for-age (**Figure 4 Panel A**) versus height-age-for-age (**Figure 4 Panel B**). In the Lulun trial, there was essentially no change in the control group’s LAZ with age (from baseline to end-line), while the intervention group’s LAZ increased towards 0 (Figure 4 Panel A), and the corresponding change in height-age was nearly parallel with the change in chronological age (Figure 4 Panel B). No major differences were visually apparent between the intervention and control groups in the Mazira trial on the LAZ-for-age or height-age-for-age scales.

**FIGURE 4.**
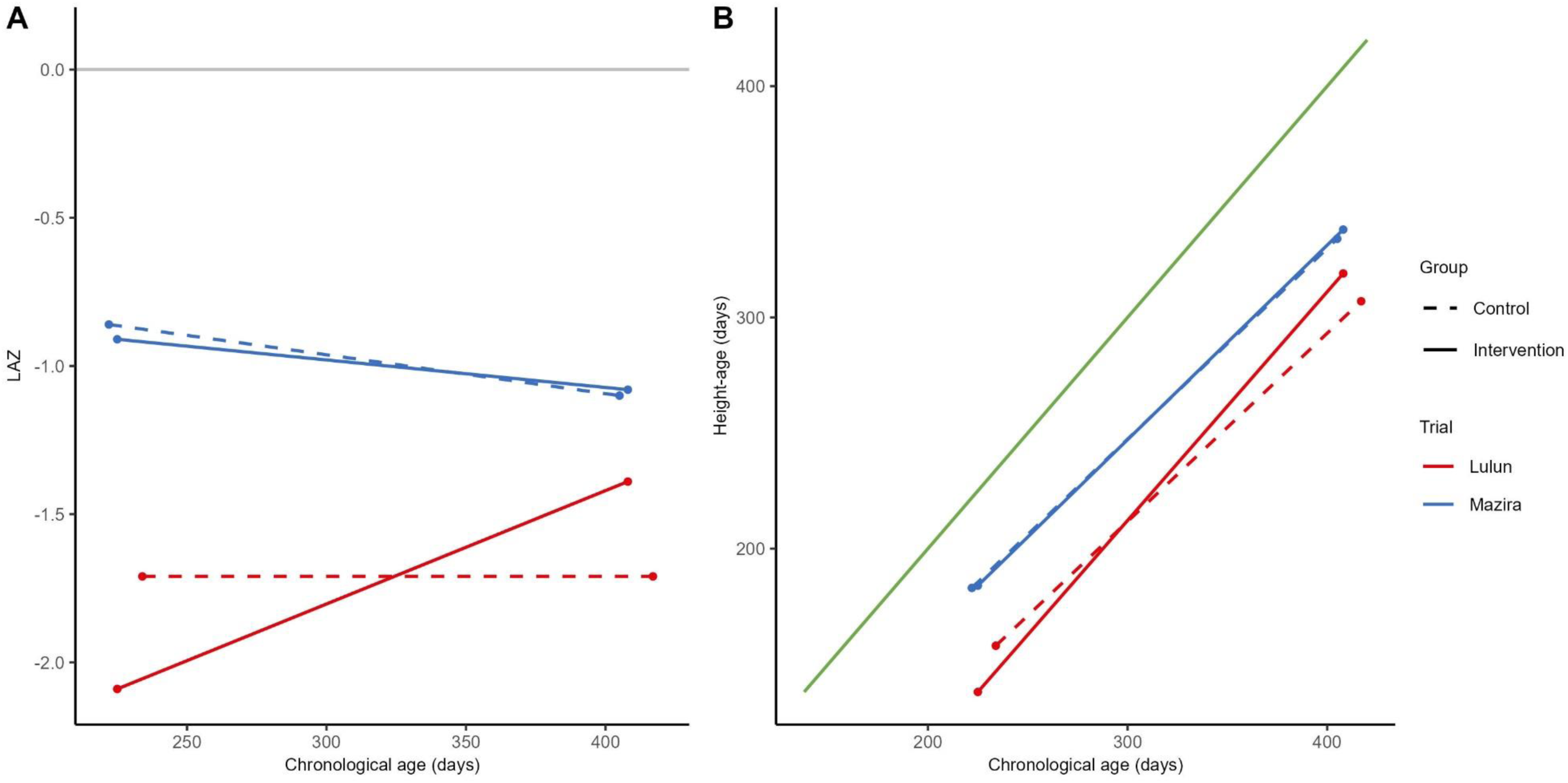
Linear growth from the start to end of the intervention period in the Lulun and Mazira egg supplementation trials expressed conventionally using mean LAZ (Panel A) and height-age (Panel B). The grey line in panel A) represents the WHO Growth Standards (WHO-GS) median (LAZ=0). The green line in panel B) represents the WHO-GS median when expressed as height-age as a function of chronological age. LAZ, length-for-age Z-score. Data sources: Iannotti LL, Lutter CK, Stewart CP, Gallegos Riofrío CA, Malo C, Reinhart G, et al. Eggs in Early Complementary Feeding and Child Growth: A Randomized Controlled Trial. J Pediatr. 2017;140(1). Stewart CP, Caswell B, Iannotti L, Lutter C, Arnold CD, Chipatala R, et al. The effect of eggs on early child growth in rural Malawi: the Mazira Project randomized controlled trial. Am J Clin Nutr. 2019;110(4):1026-33.

The sensitivity analysis showed that height-age estimates derived from mean length/height (n=2 trials) did not notably differ from estimates calculated from mean LAZ/HAZ, provided the age interval over which data was aggregated was narrow (**Supplemental Table 2**).

## DISCUSSION

Childhood linear growth faltering is a pervasive public health problem in LMICs for which progress towards global targets has been incomplete (32). New methods of communicating the results of controlled trials and program evaluations may influence public health priority-setting. In this work, height-age was used as an alternative expression of group-average child linear growth, and the PMB was used to quantify the extent of linear growth improvement achieved by nutrition-specific interventions tested in RCTs in LMICs.

Application of height-age and the PMB metric to assess the effect of interventions builds on findings from prior work demonstrating that the analysis of changes in mean HAZ with age does not enable clear determinations of group-level catch-up growth (25). Due in part to the popularity of the *Victora plots* (HAZ-by-age curves), it is conventionally assumed that stable HAZ often seen after 2 years of age (beyond the *first 1000 days*) reflects normal average growth and that an upward trend in HAZ (towards HAZ=0) is evidence of catch-up (33). However, plotting of the Lulun trial results showed that the control group’s stable HAZ was sub-optimal on the height-age-for-age scale.

Early definitions of catch-up growth suggested that the goal of catch-up was to return children to their pre-faltering growth curve (34, 35), which may have provided the basis for this assumption in HAZ-tracking. However, expecting an intervention to return a previously faltered population-average HAZ to the median for chronological age (HAZ=0) is usually unrealistic, as this would suggest that growth proceeds at a rate faster than expected for chronological and skeletal age. While this may be observed at the individual level (34), linear growth faltering in LMICs is due to numerous factors, including inadequate dietary intake, infections and other inflammatory stimuli, and psychosocial factors (2). Public health interventions, including nutrition-specific interventions such as micronutrient supplementation, are often implemented for a relatively short period of time and do not address all the factors limiting linear growth. Therefore, even small increases in HAZ (or reductions in stunting) may be unlikely, highlighting a need to temper general expectations and quantify the growth that could feasibly occur in these settings (8).

The PMB incorporates a theory-based benchmark of optimal growth that could occur in LMIC settings, based on a current understanding of the biology of long bone growth. In developing the PMB metric, we assumed that a PMB of 100% would not be a realistic expectation for a single intervention that is not designed to resolve all of the many complex factors inhibiting children’s growth in LMICs. As the PMB metric was developed and applied for the first time in this work, we were unable to define thresholds of success on the PMB scale. Even so, the PMB has potential value as it is a standardized measure of the extent to which optimal growth potential was achieved, permitting assessment of the comparative efficacy of interventions and/or programs because it is inherently normalized across ages and intervention durations. Further application of the PMB metric to a wider array of interventions previously tested in LMICs is needed to define the range of plausible PMB values and establish guidelines for using the PMB in priority-setting.

A secondary benefit of the PMB is that it may serve as a plausibility check of trial results in LMICs if a PMB of 100% is approximated or even exceeded. Although mathematically possible, a PMB>100% would imply that the intervention promoted growth beyond what is expected for children’s skeletal age, which is unlikely to occur even when substantive, permanent changes are made to children’s environments (36). Given these expectations, an observed PMB of 94%, with an upper 95% CI exceeding 100%, as calculated for the Lulun trial, should be interpreted with caution. When the Lulun trial was first reported, it received considerable attention due to the impressive apparent effect on linear growth (37, 38), especially given the simplicity of the intervention (19). However, the findings of the Lulun trial were not confirmed in a similar trial in Malawi (the Mazira study), which had a much larger sample (about 4 times greater than the Lulun trial) (20). It is possible the Lulun trial findings occurred due to imperfect randomization (19), and the very high PMB suggests the findings are unlikely to be replicable.

Findings of this study provide a toolkit for researchers to use in future applications of height-age and the PMB to RCT analysis. First, we found that in secondary use studies (i.e., re-analysis of published results), acceptable group-average height-age estimates can be derived from reported mean HAZ if mean height is not available. When IPD is available, it is feasible to generate individual child-level height-age estimates from individual-level height, and then calculate the population-average height-age. However, individual child height-age estimates are not readily interpretable as they incorrectly imply that each child should track along the median curve of the growth standard. The sensitivity analysis in the pilot study showed that derived height-age estimates are sensitive to the width of the age range over which data are aggregated. Therefore, estimating mean height from individual-level height data collected over a *narrow* age range and then deriving the corresponding mean height-age is currently the recommended approach. These and additional recommendations are summarized in **Figure 5**.

**FIGURE 5.**
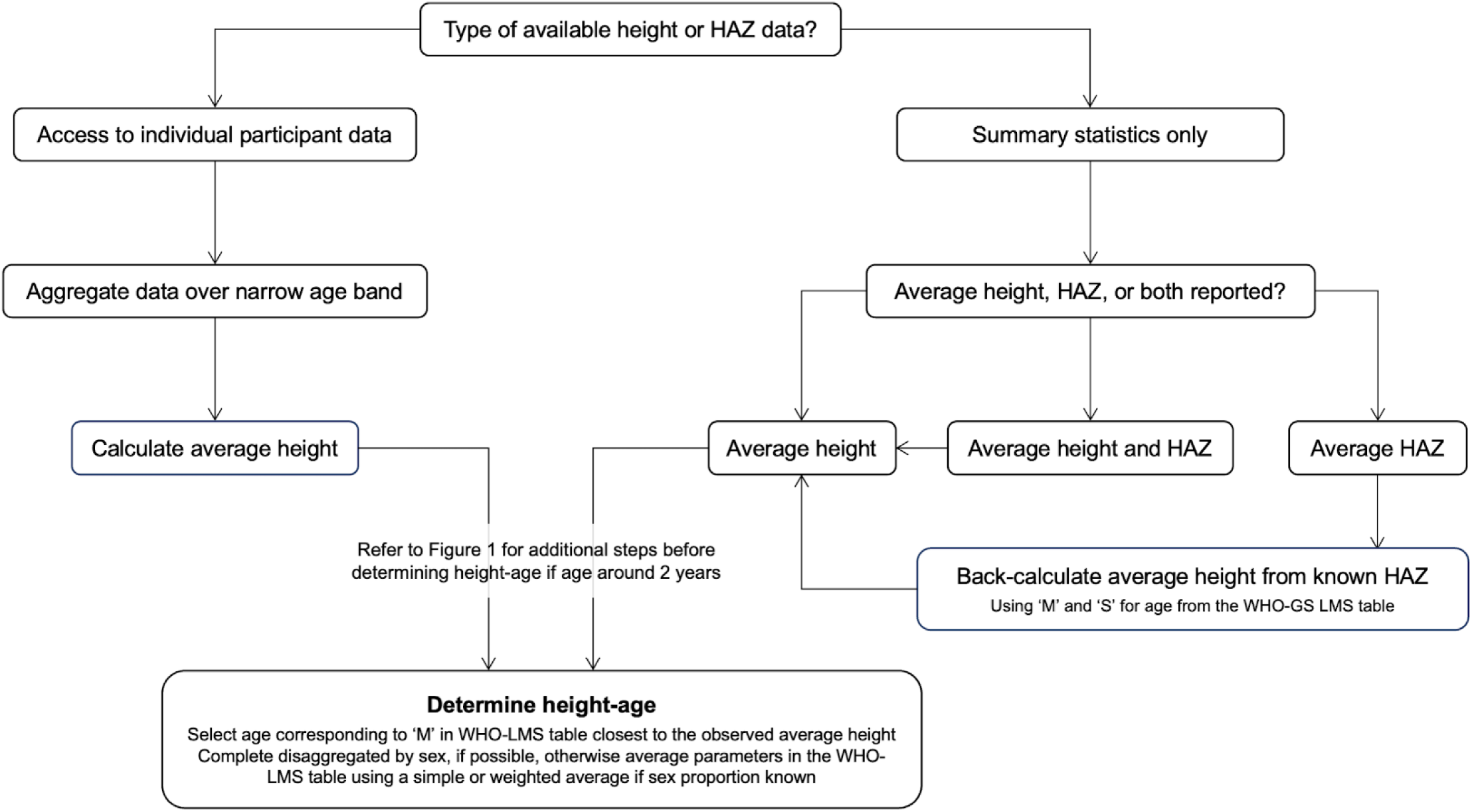
Workflow for determining height-age based on methods developed in this work. Height and HAZ are used in this figure to represent measured stature (height or length) of children. LMS, lambda-mu-sigma; M, WHO-GS median height; S, WHO-GS coefficient of variation; WHO-GS, World Health Organization Growth Standard.

Bone age based on radiographic criteria is another method of estimating skeletal age; however, height-age may be preferable in faltered populations as it might provide a more accurate portrayal of delays in long bone growth, which contributes to child height, whereas bone age is based on radiographs of smaller bones in the hand and wrist (36). Height-age is also more practical in LMICs and can be used to re-express previously collected height data in trials. However, a limitation of the PMB metric is that it has additional data requirements beyond what is needed for end-line comparisons of mean HAZ, as evidenced by the incompleteness of results in the pilot study (Table 4). Also, this study relied on the WHO-GS LMS table for children 0 to 5 years of age to select a height-age, yet growth faltering in LMICs has a prenatal onset and progresses in the early postnatal period even with exclusive breastfeeding (39), such that postnatal length deficits may be severe enough that the average observed length is smaller than the WHO-GS median length at day 0. As well, there has been increased emphasis on extending efforts for improving child growth into adolescence (i.e., beyond 5 years of age) (40, 41), so height-age methods should be further developed to encompass these age groups. Future work will incorporate other growth references, such as the INTERGROWTH-21st standards for infants (42) and the WHO growth reference data from 5 to 19 years, to extend the use of these methods from birth to adolescence.

Height-age is an alternative growth metric that can be used in place of HAZ and may offer advantages for reporting and interpreting group-average faltering and catch-up patterns in RCTs. Given the entrenched reliance on mean HAZ and stunting prevalence in global child health research, program evaluation, and country-level monitoring in LMICs, researchers may be reluctant to adopt alternative expressions of child growth. Therefore, elaboration and further applications of the height-age-based methods developed in this work are needed to expand and validate their use to stimulate widespread adoption.

## Abbreviations

BONUSKids: BONe and mUScle health in Kids
CI: confidence interval
DIVIDS: Delhi Infant Vitamin D Supplementation
HAZ: height-for-age z-score
IPD: individual participant data
LAZ: length-for-age z-score
LMICs: low– and middle-income countries
LMS: lambda-mu-sigma
MD: mean difference
MDIG: Maternal vitamin D for Infant Growth
PMB: proportion of maximal benefit
RCT: randomized controlled trial
REB: research ethics board
SD: standard deviation
SE: standard error
SickKids: The Hospital for Sick Children
WHO-GS: WHO growth standards

## Acknowledgements

DER, DGB, KMW, KMOC, ASBD, HQ, and CYC designed research; KMW conducted research; KMW, ASBD, HQ, and CYC analyzed data; and KMW and DER wrote the paper. KMW and DER had primary responsibility for final content. All authors read and approved the final manuscript.

## Data Availability

Data described in the manuscript and code for the individual participant data analysis will not be made available because this data was from a trial for which consent for public sharing of individual participant data was not sought. Study-level data described in the manuscript and code for the pilot study came from publicly available reports of trials, and can be found in the respective published report cited in this paper; the extracted dataset used in this study is also available at https://github.com/kelly-watsonn/HA1.

## Author Discolsures

The authors have no conflicts of interest or financial relationships relevant to the article to disclose.

