## Supplementary Material for "Height-age as an alternative to height-for-age z-scores to assess the effect of interventions on child linear growth in low- and middle-income countries"

### 1. Supplementary Methods

#### 1.1 Calculating measures of precision for the PMB

**Step 1**: The standard deviation (SD) of the change in height-age during the intervention period for each group, denoted as $SDC_{0:1}$, was calculated using *Equation S1* (1).

*Equation S1:* $$SDC_{0:1} = \sqrt{SD_0^2 + SD_1^2 - (2 * Correlation * SD_0 * SD_1)}$$

Variable definitions, whereby $SD_0$ represents the group's mean height-age SD at baseline, $SD_1$ represents the group's mean height-age SD at end-line, and '*Correlation'* is the Pearson correlation (Pearson product-moment correlation coefficient) between a group's baseline ('0') and end-line ('1') height-age.

**Step 1.1:** We used individual participant data (IPD) from the MDIG and BONUSKids studies to model the correlation between height-age at all possible combinations of starting and follow-up ages. This model was used to calculate a correlation value (input to Equation 1) for any study without individual-level data (provided the study population was similar to that of the MDIG trial cohort), based on children's age at baseline and end-line of the intervention period. Pearson correlation values from the length correlation matrix were used to develop the model of correlation coefficients (0, 3, 6, 9, 12, 15, 18, 21, 24 months)[1]. A simple linear regression model of the resulting Pearson correlations was used, with starting age and follow-up age as the two explanatory variables, as shown below. All analyses were performed in STATA software version 17 (STATA Corp LLC).

$$\text{Correlation} = \beta_0 + \beta_1(\text{starting age}) + \beta_2(\text{follow up age})$$

[1] It was of interest to include the birth measurement from the MDIG trial in the correlation matrix to maximize the number of observations included in the final model; however, height-age cannot be calculated at birth due to the truncation of age values in the WHO-LMS table at 0 days, which methods in this work did not address. Given that correlation coefficients are scale independent, it was expected that correlation values at all other time points would be similar between the length and height-age matrices as height-age is derived from length. This was confirmed by comparing height-age and length matrices. Thus, the length correlation matrix was used for modelling purposes.

**<u>Step 2</u>:** The $SDC_{0:1}$ and end-line sample size for each group were then used to calculate the pooled SD, standard error (SE) and 95% CI of the mean difference between group's change in height-age (from baseline), using the method for two independent samples with a continuous outcome, assuming an alpha level of 5% (2).

### 2. Supplementary Results

#### 2.1 Correlation model

The resulting model was:

Correlation = 0.797 + 0.016(starting age in months) - 0.004(follow up age in months)

Model output is presented in **Table 1**. This model was to be used to determine the correlation value to input into *Equation S4* when calculating the standard deviation of the change in height-age (from baseline to end-line), a necessary step to determine measures of precision for the novel proportion of maximal benefit (PMB) metric.

For example, if the standard deviation of the change in height-age from baseline to end-line was being calculated, and the baseline of the intervention period was 6-months, and the end-line was 12-months, the correlation value would be: 0.797 + 0.016(6) - 0.004(12) = 0.845.

**Supplementary Table 1** Linear regression output of Pearson correlation values between subsequent length measurements

| **Correlation model** | n | Regression coefficient | 95% CI | p-value |
|---|---|---|---|---|
| Starting age, months | 45 | 0.016 | 0.014, 0.018 | < 0.001 |
| Follow-up age, months | 45 | -0.004 | -0.005, -0.003 | < 0.001 |
| Intercept | 45 | 0.797 | 0.772, 0.821 | < 0.001 |

#### 2.2 Sensitivity analysis

**Supplementary Table 2** Comparison of height-age estimates determined from mean LAZ/HAZ versus length/height

| Trial (intervention duration) | Group | End-line | | | Follow-up[2,4] | | |
|---|---|---|---|---|---|---|---|
| | | | Mean height-age (95% CI), d | | | Mean height-age (95% CI), d | |
| | | *n* | Length-HA | LAZ-HA | *n* | Height-HA | HAZ-HA |
| MDIG (0-6 months)[1] | C | 206 | 153 (148, 159) | 153 (148, 158) | 107 | 1248 (1207, 1291) | 1249 (1207, 1291) |
| | I | 210 | 156 (151, 161) | 157 (152, 162) | 114 | 1266 (1234, 1299) | 1265 (1232, 1299) |
| DIVIDS (0-6 months) | C | 638 | NA[3] | 118 (116, 121) | 466 | 1347 (1309, 1385) | 1356 (1335, 1377) |
| | I | 620 | NA[3] | 122 (119, 124) | 446 | 1351 (1313, 1390) | 1370 (1349, 1392) |

[1]Different primary end-line selected for this study (6 months) than the primary end-line reported in the MDIG trial (1 year). [2]'Follow-up' represents measurement of children after an elapsed period following cessation of the trial. [3]Length data not available. [4]The average age of children at the DIVIDS trial follow-up in both groups was 5 years, with a standard deviation of 1 year.

CI, confidence interval; d, days; DIVIDS, Delhi Infant Vitamin D Supplementation; HA, height-age; HAZ, height-for-age z-score; LAZ, length-for-age z-score; MDIG, Maternal Vitamin D for Infant Growth.
